# Cardiorenal Effects of Angiotensin-converting enzyme inhibitors and Angiotensin receptor blockers in people underrepresented in trials: analysis of routinely collected data with validation against a target trial

**DOI:** 10.1101/2022.12.02.22282220

**Authors:** Paris J Baptiste, Angel YS Wong, Anna Schultze, Catherine M Clase, Clémence Leyrat, Elizabeth Williamson, Emma Powell, Johannes FE Mann, Marianne Cunnington, Koon Teo, Shrikant I Bangdiwala, Peggy Gao, Laurie Tomlinson, Kevin Wing

## Abstract

**Background:** Cardiovascular disease (CVD) is a leading cause of death globally. Angiotensin-converting enzyme inhibitors (ACEi) and angiotensin receptor blockers (ARB), compared in the ONTARGET trial, each prevent CVD. However, trial populations may not be representative of the general population.

**Methods:** Using trial replication methods within routine-care data, we explored replicability of the ONTARGET trial. For people prescribed an ACEi and/or an ARB in the UK Clinical Practice Research Datalink CPRD GOLD from 1/1/2001-31/7/2019, we applied trial criteria and propensity-score methods to create an ONTARGET trial-eligible cohort. Comparing ARB to ACEi, using Cox-proportional hazards models, we estimated hazard ratios for the primary composite trial outcome (cardiovascular death, myocardial infarction, stroke, or hospitalisation for heart failure), as well as secondary outcomes. As the pre-specified criteria were met confirming trial replicability, we then explored treatment effect heterogeneity of ACEi and ARB among three trial-underrepresented subgroups: females, those aged ≥75 years and those with chronic kidney disease.

**Findings:** In the trial-eligible population (n=137,155), results for the primary outcome met pre-specified criteria for similarity to the ONTARGET trial and demonstrated similar effects of ARB and ACEi, (HR 0.97 [95% CI: 0.93, 1.01]). When extending to trial-underrepresented groups, similar treatment effects of ARB and ACEi were observed by sex (P=0.09), age (P=0.70) and chronic kidney disease status (P=0.10).

**Interpretation:** We were able to replicate the results of the ONTARGET trial using routinely-collected healthcare data. Results suggest that trial findings were generalisable to population subgroups underrepresented in the trial.

**Funding:** GlaxoSmithKline

**Research in Context:** *Evidence before this study:* Trial replication is an important methodology increasingly used to validate findings from observational studies against target trials. Unlike many naïve observational comparisons, a previous study demonstrated replicability of the ONTARGET trial using United States insurance claims data. However, it is unknown whether trial replicability can be extended to UK routinely-collected healthcare data. In addition, little work has been done to extend findings of comparative effectiveness among trial-underrepresented subgroups such as women, the elderly and those with chronic kidney disease despite high rates of prescribing of ACE Inhibitors and angiotensin receptor blockers among these groups in routine-care.

*Added value of this study:* With access to the individual patient data from the ONTARGET study and using propensity-score methods to address confounding, we demonstrated trial replicability using routinely-collected primary care data, representative of a large proportion of the UK population. We were then able to leverage the large sample size of the trial-eligible cohort to extend findings to trial-underrepresented groups and demonstrated similar comparative effectiveness for subgroups of patients treated with ARB and ACEi among women, those aged ≥75 years and those with chronic kidney disease.

*Implications of all the available evidence:* Our findings support similar effectiveness for cardiovascular and renal outcomes for patients receiving an ARB compared to an ACEi in a trial-eligible cohort and subgroups for which there is currently a lack of evidence of treatment effectiveness. Trial-replication methodology can be used to provide evidence for populations underrepresented in clinical trials.

## INTRODUCTION

Cardiovascular disease (CVD) is a leading cause of death globally, with older people and those with chronic kidney disease (CKD) at particularly high-risk.[1] Medications used to prevent cardiovascular events are prescribed based on evidence from randomised controlled trials. However, there is uncertainty whether trial evidence is generalisable to all patient groups because trials often restrict inclusion to younger patients with fewer comorbidities,[2, 3] and because trial patients are likely to have better adherence and monitoring. Observational studies using routinely-collected healthcare data can use trial-replication methods to validate findings against those from randomised trials, sometimes referred to as “benchmarking”.[9-12] When similar findings are observed, we have more confidence that sources of bias and confounding are minimised, and aided by a large sample size and more diverse population, can then examine treatment effectiveness in trial underrepresented or excluded groups.

ONTARGET was a large global trial with 25620 participants and a follow-up of 3.5-5.5 years that compared the cardiovascular effects of angiotensin receptor II blocker (ARB) (telmisartan) to angiotensin-converting enzyme inhibitor (ACEi) (ramipril) among patients who had vascular disease or high-risk diabetes.[13, 14] Ramipril had previously been shown compared with placebo, to reduce the composite outcome of myocardial infarction, stroke or cardiovascular death by 22% (95% CI: 14-30%).[15] The findings of the ONTARGET trial of non-inferiority for telmisartan vs ramipril led to telmisartan’s licensing for cardiovascular event reduction in 2009[16] and were a major contribution to perception of equivalent treatment effectiveness for ARB and ACEI. However, the relative effectiveness of ARB and ACEI for patients not included or underrepresented in ONTARGET remains uncertain. The aims of this study were to demonstrate whether the primary and secondary outcome results of the ONTARGET trial could be replicated in UK routinely-collected data and, if so, to examine treatment effects in females, those aged ≥75 years and those with CKD, all groups that were underrepresented in ONTARGET.

## METHODS

### Data sources and study cohort

We aimed to replicate the ONTARGET trial by developing a propensity-score—weighted trial-eligible cohort in the UK Clinical Practice Research Datalink (CPRD) GOLD primary care dataset, linked to hospitalisation data from Hospital Episode Statistics (HES) and death registrations from the Office for National Statistics (ONS). We selected patients who were ever prescribed any dose of an ACEi and/or an ARB from 1/1/2001-31/7/2019 and had been registered at an up-to-standard practice (meeting minimum data quality criteria[17]) for at least 12 months at the time of their first prescription (Figure 1). We defined ‘exposed periods’ as all continuous courses of therapy, with a calculated prescription gap of >90 days referred to as an ‘unexposed period’. We did not restrict the study cohort to new users; therefore, we started their follow-up at the start of any of the exposed periods for which they met trial criteria and were included in the cohort, thus emulating recruitment into the ONTARGET trial. Patients were not required to have a minimum length of exposure to be considered. Using Read diagnostic and ICD-10 codes, we selected exposed periods that met the ONTARGET trial criteria. This resulted in a pool of trial-eligible exposed periods within individuals in CPRD. Specific diagnostic codes used for cohort identification are available for download: https://doi.org/10.17037/DATA.00002112. Our study protocol has been published;[18] Supplementary Table 1 details post-hoc changes to the published protocol.

**Figure 1.**
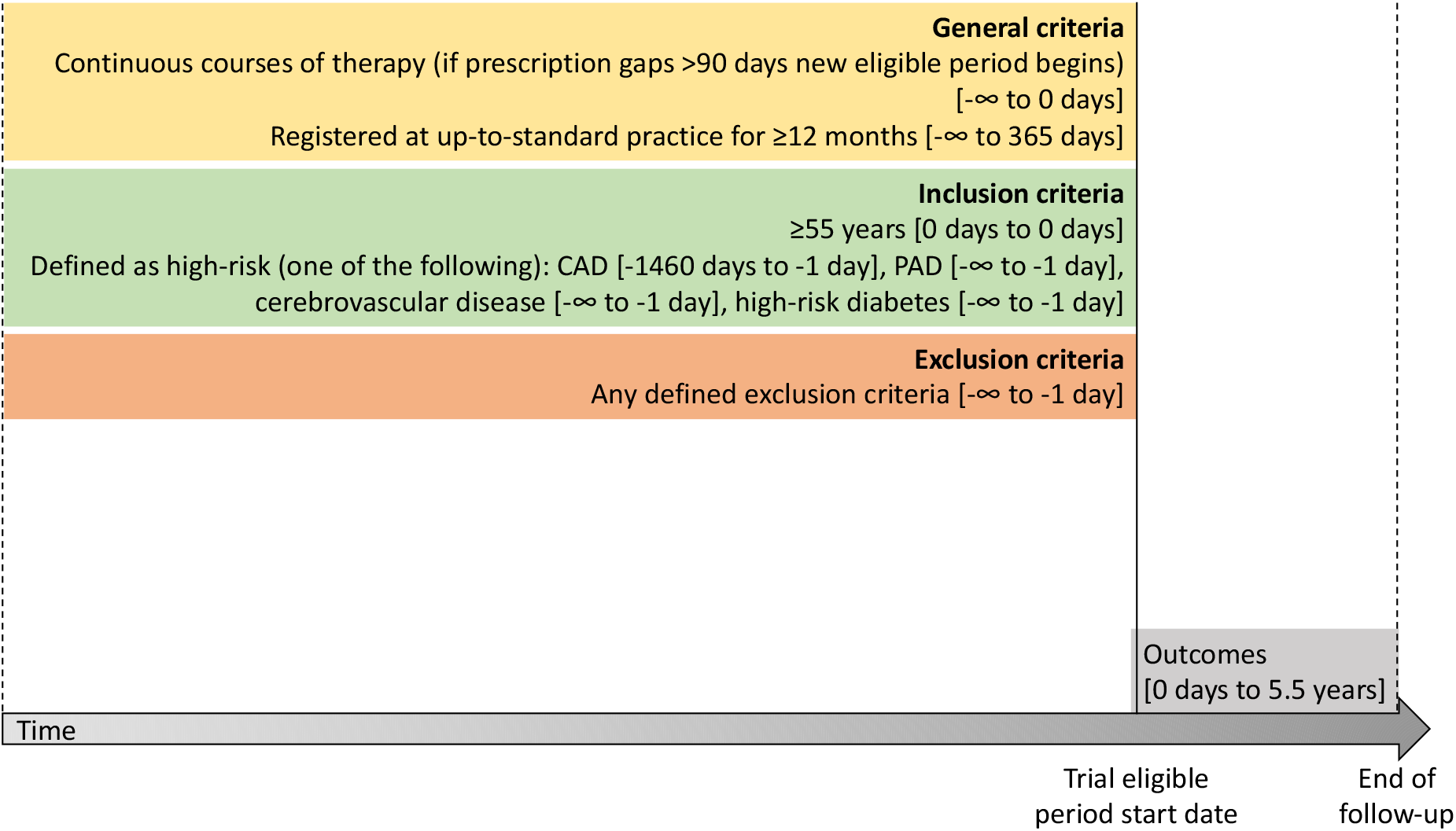
Study diagram End of follow-up was earliest of date of outcome, transferred out of practice date, death date, date of last collection, or 5.5 years from the start of eligible period. Trial eligible periods are defined as exposed periods where all trial criteria are met prior to the start of the exposed period. An exposed period is defined as periods of continuous courses of therapy (<90 days between prescriptions). CAD=coronary artery disease, PAD=peripheral artery disease. Details of how inclusion and exclusion criteria were defined are published previously. An up-to-standard practice is one that meets minimum data quality criteria based on continuity of recording and recorded number of deaths.

#### Trial replication

The original trial coordinator, Population Health Research Institute, anonymised and provided access to the ONTARGET trial data. The trial data were combined with the CPRD cohort of trial-eligible ACEi exposed periods. We then 1:1 matched each ONTARGET trial participant to one trial-eligible ACEi exposed period, without replacement, on closest propensity-score, using a propensity-score model for the probability of being included in the trial (Model 1). We used a caliper of 0.25 of the standard deviation of the logit of the propensity-score, with the restriction that only one ACEi trial-eligible exposed period per patient could be matched. This resulted in a trial-matched ACEi cohort. We used standardised differences (<0.1) and kernel density plots to assess the quality of matches.[19]

To ensure balance among CPRD groups used in analysis, we appended the trial-matched ACEi cohort with the trial-eligible ARB cohort to develop a second propensity-score model for the probability of receiving an ACEi (Model 2). To generate the propensity-score— weighted trial-eligible cohort for our main analysis, we applied Model 2 to a cohort containing one randomly selected ARB and ACEi trial-eligible period per patient, respectively, generated propensity-scores and obtained inverse probability weights.[20] Ensuring balance using propensity-score weights instead of matching, enabled us to maximise the number of participants included in the analysis. Patients could contribute to both ARB and ACEi exposed cohorts, but patients could not be matched to themselves, and trial-eligible periods that had prescription start dates on the same day were excluded from both groups.

Variables included in propensity-score Model 2 were chosen based on *a-priori* knowledge of predictors of treatment with an ACEi, and are displayed in Supplementary Table 2. To maximise comparability between the ACEi and ARB trial-eligible cohorts generated from routine data, additional variables not available in ONTARGET, such as socio-economic status, were included in Model 2. Because our cohort included prevalent users, we also included variables associated with switching treatments, such as time since first trial-eligible period, and number of previous ARB/ACEi trial-eligible periods.[21]

### Procedures

#### Exposures and outcomes

To maximise study power and generalisability, we compared outcomes between users of ARB and ACEi, rather than telmisartan and ramipril specifically. Outcomes were selected to replicate those in the ONTARGET trial.

- Primary outcome: composite of cardiovascular death, myocardial infarction (MI), stroke, or hospital admission for congestive heart failure
- Secondary outcomes:
  - Main secondary outcome: composite of cardiovascular death, MI or stroke
  - Individual components of primary outcome
  - Death from non-cardiovascular causes
  - All-cause mortality
- Further secondary and other outcomes: (separately) newly-diagnosed congestive heart failure; revascularisation procedures; loss of glomerular filtration rate (GFR) or development of end-stage kidney disease (ESKD) (defined as: 50% reduction in estimated GFR (eGFR), start of kidney replacement therapy (KRT) or development of eGFR < 15ml/min/1.73m^2^); development of ESKD (defined as: start of KRT or development of eGFR < 15ml/min/1.73m^2^); microvascular complications of diabetes mellitus. GFR was calculated using the CKD-Epi equation 2009 without reference to ethnicity.[22]
- Safety outcomes: cough; angioedema; hyperkalaemia (potassium >5.5 mmol/L); ≥30% increase in serum creatinine.

### Statistical analysis

#### Trial replication

Using an intention-to-treat approach for the main analysis, we compared cohorts using a Cox proportional hazards model weighted by propensity-scores generated from Model 2, with robust standard errors. The Cox model was additionally adjusted for any variables that demonstrated imbalance after propensity-score—weighting.[23] To replicate the trial per-protocol analysis, we also carried out an on-treatment analysis of ARB vs ACEi, additionally censoring at discontinuation of trial-eligible period. Further details on censoring are specified in Supplementary Figure 1.

Because ONTARGET reported relative risks for safety outcomes, we used a logistic regression model with robust standard errors. Treatment cessation was defined as the end of an included trial-eligible exposed period (i.e., a prescription gap of >90 days after the calculated prescription end date). The last safety event which occurred before treatment cessation was considered as the reason for treatment cessation and these results were compared with ONTARGET.

We replicated the subgroup analyses carried out in ONTARGET using a propensity-score— weighted Cox proportional hazards model fitted with an interaction term for subgroup and treatment and used a Wald test to identify any effect modification. The subgroups studied were as in ONTARGET: sex, age (<65 years, 65-74 years, ≥75 years), systolic blood pressure (SBP) (≤134 mmHg, 135-150 mmHg, >150 mmHg), diabetes, and cardiovascular disease at study entry. In addition, we included CKD status at baseline as a subgroup (CKD: eGFR <60 mL/min/1.73m^2^).

### Validation criteria

A priori, we defined replicability of the primary outcome of ONTARGET (HR 1.01 [95% CI: 0.94, 1.09]) if the HR estimates from the propensity-score—weighted analysis for ARB vs ACEi were between 0.9-1.12 and the 95% CI for the HR contained 1.0.[18]

### Underrepresented groups

Conditional on the validation criteria being met, we examined whether there was treatment heterogeneity among the underrepresented group using interaction terms for sex, age and CKD status. For CKD status, we repeated methods to create the propensity-score—weighted cohort after removing the trial exclusion criteria of baseline serum creatinine >265 μmol/L.

### Sensitivity analyses

To explore any benefits to using a propensity-score—matched trial-eligible cohort, which ensured patient characteristics were comparable to trial participants, as opposed to a propensity-score—weighted trial-eligible cohort where patients were more diverse, we 1:1 matched the trial-matched ACEi patients to closest trial-eligible ARB period using propensity-scores from Model 2 and repeated the analyses.[18]

To examine the impact of including patients who may have only received one prescription for an ARB/ACEi, we started follow-up from 28 days after the start of the trial-eligible period, excluding patients if there were no prescriptions after 28 days.

We assessed the impact on the kidney outcomes of specifying sustained deterioration of kidney function. This required eGFR <15ml/min/1.73m^2^ or 50% reduction in eGFR on two occasions at least 3 months apart for loss of eGFR or ESKD and development of ESKD outcomes.

As a post-hoc sensitivity analysis, we assessed the impact of changing between medications in CPRD for safety outcomes by restricting the cohort to patients’ first trial-eligible exposed period, and by excluding those with previous exposure to the alternative drug at any time before.

## Role of the funding source

This project is funded by a GlaxoSmithKline PhD studentship as part of a collaboration between GlaxoSmithKline and the London School of Hygiene and Tropical Medicine. The funders had no role in the study design, data collection, data analysis, data interpretation, or writing of the report.

## Results

### Baseline characteristics

After propensity-score—weighting, 96,602 ACEi and 40,553 ARB prescribed patients were included in the comparison of ARB vs ACEi users (Supplementary Figure 1).

Mean age was similar across exposure groups (71 years), slightly older than in ONTARGET (66 years). There was a higher proportion of females across each exposure group (∼51%) than in ONTARGET (27%) (Table 1). Balance before and after weighting is shown in Supplementary Table 3. Imbalance remained for several time-related variables: time since first trial-eligible period, calendar year of trial-eligible period and number of prior ARB trial-eligible periods, so analysis was adjusted for these variables.

**Table 1.** Baseline characteristics of trial-eligible patients after applying trial criteria included in propensity-score—weighted analysis compared to ONTARGET

|  |  |  |  |
| --- | --- | --- | --- |
| **Characteristic** | **ACEi**<br>N=96,602 | **ARB**<br>N=40,553 | **ONTARGET**<br>N=25,620 |
| **Age - year** | 70.8 ± 9.0 | 71.2 ± 8.7 | 66.4 ± 7.2 |
| **Blood pressure – mmHg** | 147.4 ± 20.7 / 80.1 ± 10.7 | 148.1 ± 20.7 / 79.7 ± 10.5 | 141.8 ± 17.4 / 82.1 ± 10.4 |
| **Body-mass index** | 28.3 ± 5.3 | 28.8 ± 5.4 | 28.2 ± 4.7 |
| **Cholesterol – mmol/l** | 4.8 ± 1.2 | 4.7 ± 1.1 | 4.9 ± 1.2 |
| **Triglycerides – mmol/l** | 1.7 ± 1.0 | 1.6 ± 0.9 | 1.7 ± 1.1 |
| **Glucose – mmol/l** | 6.5 ± 2.5 | 6.4 ± 2.4 | 6.7 ± 2.6 |
| **Creatinine - μmol/l** | 93.9 ± 25.0 | 94.3 ± 26.9 | 94.2 ± 24.4 |
| **Potassium – mmol/l** | 4.4 ± 0.5 | 4.4 ± 0.5 | 4.4 ± 0.4 |
| **Female sex – no. (%)** | 45508 (47.1) | 22690 (56.0) | 6831 (26.7) |
| **Ethnic group – no. (%)** |  |  |  |
| Black | 1280 (1.3) | 736 (1.8) | 629 (2.5) |
| Other | 1134 (1.2) | 736 (1.8) | 4901 (19.1) |
| South Asian | 3026 (3.1) | 1799 (4.4) | 1375 (5.4) |
| Unknown | - | - | 7 (<0.1) |
| White | 91162 (94.4) | 37411 (92.3) | 18708 (73.0) |
| **Clinical history – no. (%)** |  |  |  |
| CAD<sup>1</sup> | 68009 (70.4) | 28202 (69.5) | 19102 (74.6) |
| MI | 21997 (22.8) | 7301 (18.0) | 12549 (49.0) |
| Angina pectoris | 31595 (32.7) | 13205 (32.6) | 11505 (44.9) |
| Cerebrovascular disease<sup>2</sup> | 8695 (9.0) | 3140 (7.7) | 5342 (20.9) |
| PAD<sup>3</sup> | 9999 (10.4) | 4078 (10.1) | 3468 (13.5) |
| Diabetes | 43751 (45.3) | 20003 (49.3) | 9612 (37.5) |
| High-risk diabetes<sup>4</sup> | 30736 (31.8) | 14757 (36.4) | 7151 (27.9) |
| **Previous procedures – no. (%)** |  |  |  |
| CABG | 6747 (7.0) | 2912 (7.2) | 5675 (22.2) |

| Characteristic | ACEi<br>N=96,602 | ARB<br>N=40,553 | ONTARGET<br>N=25,620 |
| --- | --- | --- | --- |
| PTCA | 10055 (10.4) | 3823 (9.4) | 7437 (29.0) |
| CKD (eGFR<60) | 27608 (28.6) | 13246 (32.7) | 5470 (21.4) |
| **Smoking status – no. (%)** |  |  |  |
| Non-smoker | 34503 (35.7) | 16470 (40.6) | 9088 (35.5) |
| Current smoker | 12921 (13.4) | 3552 (8.8) | 3225 (12.6) |
| Past smoker | 49178 (50.9) | 20531 (50.6) | 13276 (51.8) |
| Unknown | - | - | 31 (0.1) |
| **Alcohol status – no. (%)** |  |  |  |
| Drinker | 76756 (79.5) | 31840 (78.5) | 10345 (40.4) |
| Non-drinker | 19846 (20.5) | 8713 (21.5) | 15261 (59.6) |
| Unknown | - | - | 14 (<0.1) |
| **Medication<sup>5</sup> – no. (%)** |  |  |  |
| ACE inhibitor | 78287 (81.0) | 4659 (11.5) | 14750 (57.6) |
| Alpha-blocker | 6892 (7.1) | 3202 (7.9) | 1095 (4.3) |
| Oral anticoagulant agent | 4613 (4.8) | 1282 (3.2) | 1939 (7.6) |
| Antiplatelet agent | 12334 (12.8) | 2482 (6.1) | 2824 (11.0) |
| ARB | 440 (0.5) | 34579 (85.3) | 2213 (8.6) |
| Aspirin | 44011 (45.6) | 9325 (23.0) | 19403 (75.7) |
| Beta-blocker | 34178 (35.4) | 6756 (16.7) | 14583 (56.9) |
| Calcium-channel blocker | 28820 (29.8) | 8515 (21.0) | 8472 (33.1) |
| Digoxin | 2533 (2.6) | 600 (1.5) | 865 (3.4) |
| Diuretics | 32002 (33.1) | 8838 (21.8) | 7164 (28.0) |
| Diabetic treatment | 20060 (20.8) | 4910 (12.1) | 8056 (31.4) |
| Nitrates | 14862 (15.4) | 3172 (7.8) | 7523 (29.4) |
| Statins | 52925 (54.8) | 11474 (28.3) | 15783 (61.6) |

N= number of patients; no. (%)=number (percent); CAD=coronary artery disease; MI=myocardial infarction; PAD=peripheral artery disease; CABG=coronary artery bypass graft; PTCA=percutaneous transient coronary angioplasty; CKD=chronic kidney disease (eGFR<60mmol/L)
One third of ONTARGET participants received both ramipril plus telmisartan.
<sup>1</sup> Includes diagnosis of: MI at least 2 days prior, angina at least 30 days prior, angioplasty at least 30 days prior, CABG at least 4 years prior
<sup>2</sup> Includes diagnosis of: stroke/TIA
<sup>3</sup> Includes diagnosis of: limb bypass surgery, limb/foot amputation, intermittent claudication
<sup>4</sup> Includes DM with: retinopathy, neuropathy, chronic kidney disease, proteinuria or other complication
<sup>5</sup> Within 3 months prior to eligible start date. Antiplatelet agent= clopidogrel/ticlopidine

### Follow-up and adherence

Among the propensity-score—weighted trial-eligible cohort, a total of 82,121 patients were followed until an event or 5.5 years of follow-up (maximum follow-up in the ONTARGET trial). After one year, among patients in the ARB group 2.6% had switched to an ACEi and among patients in the ACEi group, 11% switched to an ARB. Adherence was lower in CPRD, with 70% ACEi patients still on ACEi treatment after one year and 78% ARB patients still on ARB treatment after one year, compared to ONTARGET, where 92% ramipril patients were taking an ACEi and 94% telmisartan patients taking an ARB after one year[13] (Supplementary Table 4).

### Trial replication

#### Primary outcomes and validation

Among the propensity-score—weighted trial-eligible cohort, the primary composite outcome occurred in 6287 (16%) in the ARB group and in 16935 (18%) in the ACEi group (median follow-up 4.7 years), for event rates of 4.2 and 4.4 per 100 person-years, respectively. In ONTARGET, the number of events was 1423 (17%) and 1412 (17%) in the telmisartan and ramipril treatment groups, respectively, over median follow-up of 4.7 years. Comparing ARB users with ACEi users in the trial-eligible cohorts, the risk of the primary outcome was similar, HR 0.98 (95% CI: 0.94, 1.02) in the propensity-score–weighted, adjusted analysis. This was comparable to the ONTARGET primary outcome (HR 1.01 [95% CI: 0.94, 1.09]) and met the pre-specified validation criteria of trial replicability (Table 2 and Figure 2). Results of the on-treatment analysis were similar HR 1.02 (95% CI: 0.93, 1.12) for ARB vs ACEi, comparable to both the ONTARGET per-protocol analysis (HR 1.00 [95% CI: 0.92, 1.09]) and to our main intention-to-treat analysis.

**Table 2.** Number of events for the primary outcome, its components, and death from any cause for a propensity-score—weighted analysis of ARB vs ACEi using CPRD data.

| Outcome | CPRD |  |  | ONTARGET |
| --- | --- | --- | --- | --- |
|  | ACEi<br>(N=96,602) | ARB<br>(N=40,553) | ARB vs ACEi<br>(N=137,155) | Telmisartan vs<br>ramipril<br>(N=17,118) |
|  | *Number (percent)* |  | *Hazard ratio (95% CI)* |  |
| Primary composite: Death from cardiovascular causes, myocardial infarction, stroke, or hospitalisation for heart failure | 16935 (17.5) | 6287 (15.5) | 0.98 (0.94, 1.02) | 1.01 (0.94, 1.09) |
| Main secondary outcome: Death from cardiovascular causes, myocardial infarction or stroke | 5363 (13.2) | 14647 (15.2) | 0.98 (0.94, 1.02) | 0.99 (0.91, 1.07) |
| Myocardial infarction | 11617 (12.0) | 4090 (10.1) | 0.97 (0.92, 1.01) | 1.07 (0.94, 1.22) |
| Stroke | 3768 (3.9) | 1573 (3.9) | 1.04 (0.97, 1.12) | 0.91 (0.79, 1.05) |
| Hospitalisation for heart failure | 4028 (4.2) | 1570 (3.9) | 0.97 (0.90, 1.05) | 1.12 (0.97, 1.29) |
| Death from cardiovascular causes | 5194 (5.4) | 1825 (4.5) | 0.96 (0.90, 1.03) | 1.00 (0.89, 1.12) |
| Death from non-cardiovascular causes | 6984 (7.2) | 2649 (6.5) | 0.97 (0.92, 1.02) | 0.96 (0.83, 1.10) |
| Death from any cause | 12178 (12.6) | 4474 (11.0) | 0.97 (0.93, 1.01) | 0.98 (0.90, 1.07) |
| Notes: CPRD weighted analysis includes 1 randomly selected trial-eligible period per patient. Propensity-score—weighted with robust standard errors. Analysis adjusted for time since first eligible period, number of prior ARB periods and calendar year.<br>Myocardial infarction and stroke include both fatal and non-fatal events.<br>55,015 (57.0%) of ACEi patients included received ramipril as the first prescription for the included trial-eligible exposed period. 1,495 (3.7%) of ARB patients included received telmisartan as the first prescription for the included trial-eligible exposed period.<br>ONTARGET results are from published findings. |  |  |  |  |

**Figure 2.**
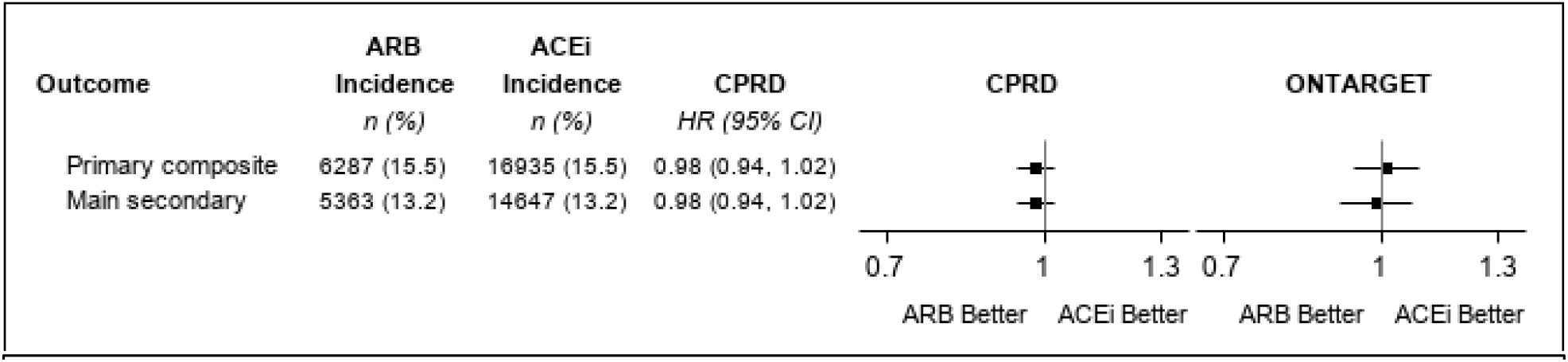
Hazard ratios for the propensity-score—weighted and adjusted analysis of ARB vs ACEi for the primary composite outcome and main secondary outcome compared to comparison of telmisartan vs ramipril in ONTARGET. n (%)= number of events (percent). Primary composite outcome: cardiovascular death, myocardial infarction, stroke or hospitalisation for heart failure. Main secondary composite outcome: cardiovascular death, myocardial infarction or stroke.

#### Secondary and other outcomes

Results were consistent with ONTARGET for the main secondary composite outcome of cardiovascular death, MI or stroke (Figure 2) and all other secondary outcomes, including development of ESKD (HR 1.06 [95% CI: 0.95, 1.19]) (Supplementary Table 5). However, within the CPRD trial-eligible cohort the risk of the composite of loss of GFR or ESKD was higher for ARB users than for ACEi users (HR 1.11 [95% CI: 1.04, 1.19]), where ONTARGET observed similar treatment effects (Table 2 and Supplementary Table 5).

#### Safety outcomes

In analyses of safety outcomes as reason for treatment cessation, cough was more common in ARB than in ACEi users (RR 1.29 [95% CI: 1.16, 1.43]) and angioedema was similar between groups, both in contrast with ONTARGET findings of reduced risk of cough and angioedema with ARB vs ACEi, however the number of events in our analysis was low, and our assessment was based on timing, whereas the ONTARGET reason for discontinuation was prospectively documented. Hyperkalaemia and ≥30% increase in serum creatinine were also more common in ARB users than in ACEi users RR 1.12 (95% CI: 1.06, 1.18) and RR 1.38 (95% CI: 1.34, 1.43), respectively (Supplementary Table 6).

#### Subgroup analysis

Results of the primary outcome for ARB vs ACEi, stratified within the same subgroups as ONTARGET are shown in Figure 3. We observed evidence of effect modification by baseline SBP (P<0.01) with a lower risk among ARB users compared to ACEi in those with baseline SBP ≤134mmHg. All other subgroups studied showed no strong evidence of treatment heterogeneity between groups, which was consistent with the findings in ONTARGET.

**Figure 3.**
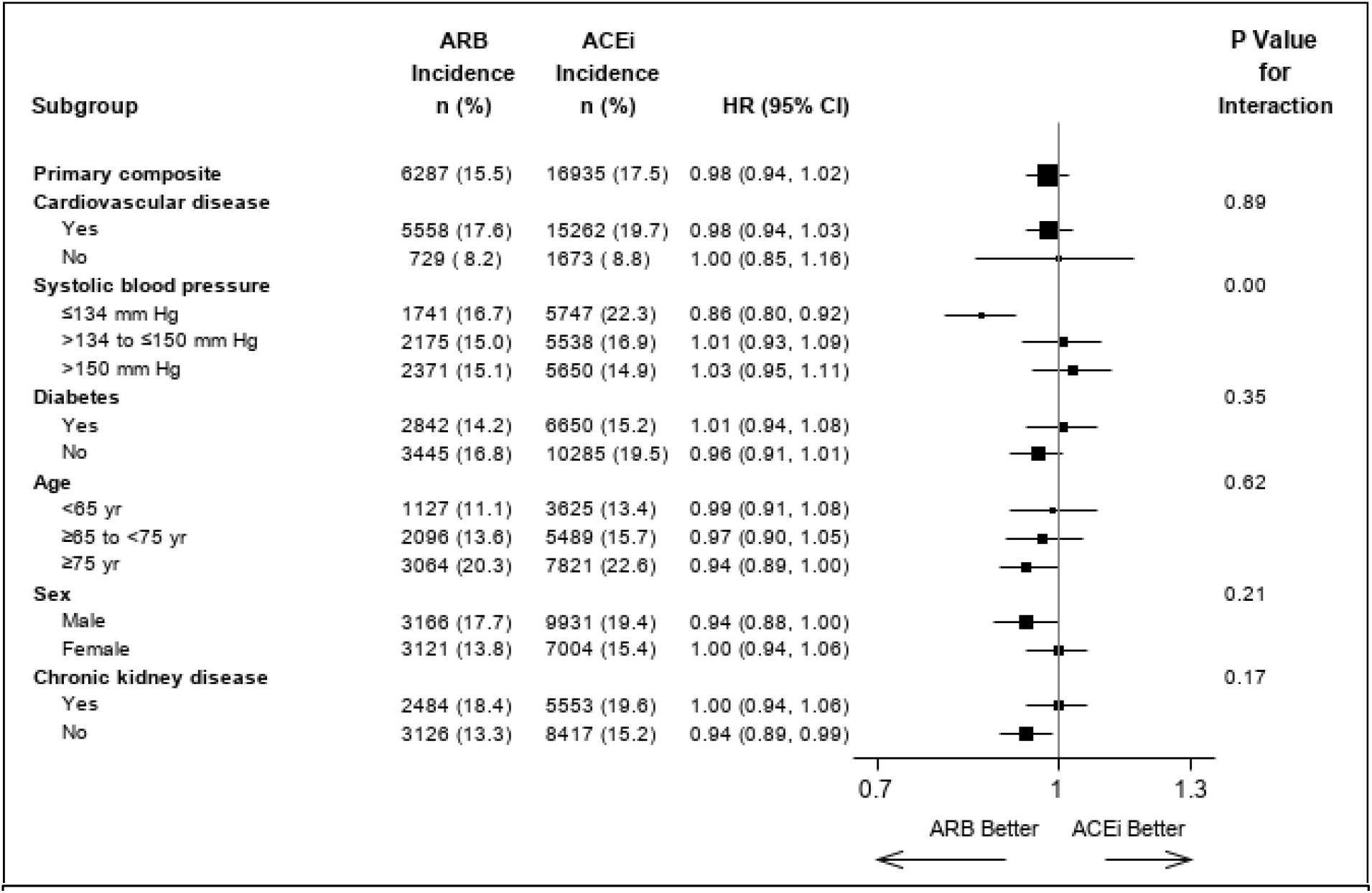
Hazard ratios in prespecified subgroups that were studied in ONTARGET (including underrepresented groups of females and aged ≥75 years) along with those with chronic kidney disease (not analysed in ONTARGET and underrepresented), for comparison of ARB vs ACEi for analysis of the primary composite outcome. n (%)= number of events (percent). P-value is the test of interaction between the treatment and each subgroup. Cardiovascular disease consists of patients with coronary artery disease, peripheral artery disease or cerebrovascular disease. Chronic kidney disease is defined as patients with eGFR<60 mL/min.

### Underrepresented groups

For ARB vs ACEi for the primary composite outcome, there was no evidence to suggest treatment heterogeneity between males and females (P=0.21), by age group (P=0.62) and by CKD status (P=0.17). Among the trial-underrepresented groups of females, those aged ≥75 years and those with CKD, treatment effects were consistent with ONTARGET (Figure 3). For most secondary outcomes treatment effects were similar among males and females. However, there was some evidence of treatment heterogeneity for the outcomes of cardiovascular-related death (P=0.03), all-cause mortality (P=0.04) and revascularisation procedures (P=0.03). Treatment effects were similar among ARB and ACEi users for men but among females ARB were associated with a lower risk of cardiovascular-related death and all-cause mortality compared to ACEi (Supplementary Figure 2).

Similarly, by age group there was no evidence of treatment heterogeneity for most secondary outcomes. However, treatment effects differed for the outcomes of revascularisation procedures (P=0.02) and loss of GFR or ESKD (P<0.01). ARB and ACEi had similar treatment effects among users aged ≥65 years but among users aged <65 years, ARB were associated with a lower risk of revascularisation procedures and a higher risk of loss of GFR or ESKD, but event numbers were low (Supplementary Figure 3).

For CKD, evidence of treatment heterogeneity was observed for MI (P=0.02) and newly diagnosed heart failure (P=0.03) and revascularisation procedures (P=0.01). For these outcomes, treatment effectiveness was similar among ARB and ACEi users with CKD at baseline but ARB were associated with a lower risk among those without CKD at baseline (Supplementary Figure 4).

### Sensitivity analyses

Analysis of the propensity-score—matched trial-eligible cohort for ARB vs ACEi gave similar results to the propensity-score—weighted trial-eligible cohort for the primary outcome (HR 0.97 [95% CI: 0.92, 1.02], number of events: ARB=2453 (16%), ACEi=2539 (16%)) (Table 2 and Supplementary Table 7). For all other outcomes, results had HRs close to 1.0 and 95% CI containing 1.0 (Supplementary Table 7).

The risk of the primary outcome was lower among ARB users when follow-up was started from 28 days after the start of the trial-eligible period (HR 0.93 [95% CI: 0.90, 0.96], number of events: ARB=5966 (15%), ACEi=16051 (16.8%)).

Specifying sustained deterioration of kidney function for loss of GFR or ESKD had no effect on results. However, among ARB users the risk of development of ESKD was increased (HR 1.16 [95% CI: 1.02, 1.32], number of events: ARB=626 (1.7%), ACEi=1016 (1.2%)). Restricting to new users with no previous exposure to the opposite drug for safety outcomes, showed a lower risk of cough and angioedema as reason for treatment cessation for ARB vs ACEi, which was consistent with the trial findings (Supplementary Table 8).

## Discussion

We emulated the ONTARGET randomised trial, using a large routinely-collected healthcare dataset. By applying the trial criteria and creating a propensity-score—weighted trial-eligible cohort with balanced characteristics in each treatment arm, we showed similar risks among ARB and ACEi users for the composite of cardiovascular death, MI, stroke or hospital admission for congestive heart failure, as well as further secondary outcomes. The ONTARGET per-protocol analysis was also replicated using an on-treatment approach where we obtained consistent results. Furthermore, marked similarity between ONTARGET and our observational study was also found in subgroup analysis, with ARB users with the lowest baseline SBP at lower risk of the primary composite outcome compared to ACEi users. We subsequently extended analysis to females, those aged ≥75 years and patients with CKD (all underrepresented in ONTARGET), where we demonstrated consistency of treatment effects for most outcomes.

### Comparison to other studies

Our findings of similar effectiveness of ARB vs ACEi by sex and age were consistent with previous comparative effectiveness studies.[24, 25] In line with the findings from a large Taiwanese cohort study,[26] we demonstrated no difference between ARB vs ACEi in risk of kidney outcomes among those with and without CKD.

One recent ONTARGET replication study using United States insurance claims data performed a propensity-score—matched analysis of telmisartan vs ramipril and found HR 0.99 (95% CI: 0.85, 1.14) for the primary outcome. [11] The sample was small (9930 patients) and, unlike the trial, included new users only.

In contrast to other naïve observational studies that have shown a decreased risk among ARB users,[27-29] we observed equal treatment effectiveness of ARB and ACEi. This implies that using trial emulation techniques and propensity-score—weighting to obtain balance among exposure groups can adequately address confounding and bias and lead to results comparable to the target trial.

### Strengths and Limitations

We were able to demonstrate that both a propensity-score—weighting approach and a propensity-score—matching approach yielded equivalent results to ONTARGET, providing evidence to support the use of a weighted approach in future trial replication studies (preferred, because weighting minimises the loss of participants involved in matching and enables greater power for examining rare outcomes such as ESKD). Having replicated the ONTARGET results, the increased sample size and diverse population in the propensity-score—weighted trial-eligible cohort allowed us to extend our analyses to trial-underrepresented groups. This included people with CKD where evidence from observational studies is limited. Among this group, we observed similar treatment effectiveness among ARB and ACEi users for the primary outcome and all other outcomes, including the outcomes of loss of GFR or ESKD and development of ESKD.

Despite overall similarity between ARB and ACEi users for most outcomes, we noted some discrepancies with ONTARGET. In the ONTARGET trial, ARB and ACEi users had comparable risk of kidney-related outcomes. In contrast, we found ARBs to be associated with a moderately greater risk of loss of GFR or ESKD compared with users of ACEi. This may reflect testing multiple outcomes, low numbers of outcomes in some strata or residual confounding by indication.

When dealing with comparisons between a new drug and a historic drug, careful considerations need to be given to handle treatment switchers and appropriately account for time trends in prescribing. We sought to account for such variables, including them as terms in our propensity-score model but it is not possible to exclude this as a source of residual confounding or bias.

Discrepancy of safety outcomes is likely due to the close monitoring of adverse events in a trial setting compared with routine clinical care. Events such as cough are likely to be underreported in routine data. In addition to this, some confounding by indication may be present, particularly for patients with a history of cough or angioedema who may have been switched from an ACEi to an ARB. This was demonstrated in our sensitivity analysis, restricting the cohort to non-switchers, where we obtained results much closer to the ONTARGET trial. However, since ARB users who have not previously been exposed to an ACEi are likely to be healthier and less likely to experience cough, due to the known risk of cough among ACEi users, we cannot be sure that restricting to non-switchers does not introduce further bias.

## Conclusion

In this emulation of the ONTARGET randomised trial using routinely-collected healthcare data, we closely replicated the primary and secondary outcomes and were able to demonstrate the generalisability of trial results to a cohort representative of patients receiving prescriptions for ACEi or ARB in UK primary care. Subsequently we were able to provide evidence that trial results extend to trial-underrepresented subgroups where evidence is limited including females, those aged ≥75 years and patients with CKD. Benchmarking findings from observational studies against target trial results can add confidence to findings when using routinely-collected data to investigate the generalisability of trial findings to wider populations.

## Data Availability

Data was provided by CPRD under terms of the LSHTM license and can be requested from CPRD. ONTARGET trial data was obtained via a data sharing agreement with the sponsor, Hamilton Health Sciences Corporation.

https://www.cprd.com/

## Acknowledgments

We are grateful to the Population Health Research Institute for providing access to the individual patient level data. This work was supported by the funding from a GSK studentship held by PB as part of an ongoing collaboration between GSK and the London School of Hygiene and Tropical Medicine.

## Author contributions

All authors contributed to the conceptualisation, study question and design. PB, LT, KW, EW, AW, AS, CL, CC and EP contributed to the statistical analysis decisions of this study. PB completed formal analysis of this study and wrote the original draft of this manuscript under the supervision of LT, KW and MC. LD facilitated the transfer of trial data used in this project. PB, LT, KW, EW, AW, AS, CL, EP had access to the data and data reported in this manuscript was verified by LT and KW. All authors contributed to the writing, review and editing of the submitted manuscript.

## Conflicts of interest

PB is funded by a GSK PhD studentship. AS is employed by LSHTM on a fellowship sponsored by GSK. CC has received consultation, advisory board membership or research funding from the Ontario Ministry of Health, Sanofi, Pfizer, Leo Pharma, Astellas, Janssen, Amgen, Boehringer-Ingelheim and Baxter. In 2018 CC co-chaired a KDIGO potassium controversies conference sponsored at arm’s length by Fresenius Medical Care, AstraZeneca, Vifor Fresenius Medical Care, Relypsa, Bayer HealthCare and Boehringer Ingelheim. CC co-chairs the cloth mask knowledge exchange, a stakeholder group that includes cloth mask manufacturers and fabric distributors. MC was an employee of GSK at the time of the study. All other authors have no conflicts.

## Notes

### Clinical Protocols

https://doi.org/10.1136/bmjopen-2021-051907

### Author Declarations

An application for scientific approval related to the use of the Clinical Practice Research Datalink (CPRD) data has been approved by the Independent Scientific Advisory Committee of the Medicines and Healthcare Products Regulatory Agency in January 2020 (protocol no. 20_012). CPRD are already approved via a National Research Ethics Committee for purely non-interventional research if this type. Ethical approval for this study has been obtained from the London School of Hygiene & Tropical Medicine Ethics Committee (Ref. 22658). Access to the individual patient data from the ONTARGET trial has been obtained by the trial investigators via a data sharing agreement.

## References

1. Jankowski, J., et al., Cardiovascular Disease in Chronic Kidney Disease: Pathophysiological Insights and Therapeutic Options. Circulation, 2021. 143(11): p. 1157–1172.

2. Maini, R., et al., Persistent Underrepresentation of Kidney Disease in Randomized, Controlled Trials of Cardiovascular Disease in the Contemporary Era. J Am Soc Nephrol, 2018. 29(12): p. 2782–2786.

3. Lee, J.S., et al., Effect of age, sex, and morbidity count on trial attrition: meta-analysis of individual participant level data from phase 3/4 industry funded clinical trials. BMJMED, 2022. 1(1).

4. Cagnacci, A. and M. Venier, The Controversial History of Hormone Replacement Therapy. Medicina (Kaunas), 2019. 55(9).

5. Rossouw, J.E., et al., Risks and benefits of estrogen plus progestin in healthy postmenopausal women: principal results From the Women’s Health Initiative randomized controlled trial. JAMA, 2002. 288(3): p. 321–33.

6. Dave, C.V., et al., Sodium-Glucose Cotransporter-2 Inhibitors and the Risk for Severe Urinary Tract Infections: A Population-Based Cohort Study. Ann Intern Med, 2019. 171(4): p. 248–256.

7. Cooper, B.A., et al., A randomized, controlled trial of early versus late initiation of dialysis. N Engl J Med, 2010. 363(7): p. 609–19.

8. Fu, E.L., et al., Timing of dialysis initiation to reduce mortality and cardiovascular events in advanced chronic kidney disease: nationwide cohort study. BMJ, 2021. 375: p. e066306.

9. Franklin, J.M., et al., Emulating Randomized Clinical Trials With Nonrandomized Real-World Evidence Studies: First Results From the RCT DUPLICATE Initiative. Circulation, 2021. 143(10): p. 1002–1013.

10. Wing, K., et al., Real world effects of COPD medications: a cohort study with validation against results from randomised controlled trials. Eur Respir J, 2021. 57(3).

11. Fralick, M., et al., Use of Health Care Databases to Support Supplemental Indications of Approved Medications. JAMA Intern Med., 2018. 178(1): p. 55–63.

12. Dahabreh, I.J., J.M. Robins, and M.A. Hernan, Benchmarking Observational Methods by Comparing Randomized Trials and Their Emulations. Epidemiology, 2020. 31(5): p. 614–619.

13. Ontarget Investigators, et al., Telmisartan, ramipril, or both in patients at high risk for vascular events. N Engl J Med, 2008. 358(15): p. 1547–59.

14. Mann, J.F.E., et al., Renal outcomes with telmisartan, ramipril, or both, in people at high vascular risk (the ONTARGET study): a multicentre, randomised, double-blind, controlled trial. Lancet, 2008. 372(9638): p. 547–53.

15. Heart Outcomes Prevention Evaluation Study, I., et al., Effects of an angiotensin-converting-enzyme inhibitor, ramipril, on cardiovascular events in high-risk patients. N Engl J Med, 2000. 342(3): p. 145–53.

16. European Medicines Agency. Micardis. 2009 25 Feb 2022; Available from: https://www.ema.europa.eu/en/medicines/human/EPAR/micardis.

17. Herrett, E., et al., Data Resource Profile: Clinical Practice Research Datalink (CPRD). Int J Epidemiol, 2015. 44(3): p. 827–36.

18. Baptiste, P.J., et al., Effects of ACE inhibitors and angiotensin receptor blockers: protocol for a UK cohort study using routinely collected electronic health records with validation against the ONTARGET trial. BMJ Open, 2022. 12(3): p. e051907.

19. Garrido, M.M., et al., Methods for constructing and assessing propensity scores. Health Serv Res, 2014. 49(5): p. 1701–20.

20. Chesnaye, N.C., et al., An introduction to inverse probability of treatment weighting in observational research. Clinical Kidney Journal, 2021: p. 1–7.

21. Suissa, S., E.E. Moodie, and S. Dell’Aniello, Prevalent new-user cohort designs for comparative drug effect studies by time-conditional propensity scores. Pharmacoepidemiol Drug Saf, 2017. 26(4): p. 459–468.

22. Levey, A.S., et al., A new equation to estimate glomerular filtration rate. Ann Intern Med, 2009. 150(9): p. 604–12.

23. Stürmer, T., et al., Analytic strategies to adjust confounding using exposure propensity scores and disease risk scores: nonsteroidal antiinflammatory drugs and short-term mortality in the elderly. Am J Epidemiol, 2005. 161(9): p. 891–8.

24. Chien, S.C., et al., Comparative Effectiveness of Angiotensin-Converting Enzyme Inhibitors and Angiotensin II Receptor Blockers in Terms of Major Cardiovascular Disease Outcomes in Elderly Patients: A Nationwide Population-Based Cohort Study. Medicine (Baltimore), 2015. 94(43): p. e1751.

25. Oger, E., et al., Effectiveness of angiotensin-converting enzyme inhibitors and angiotensin receptor blockers on total and cardiovascular mortality and morbidity in primary prevention: A nationwide study based on French Health Insurance Data (SNDS). J Clin Hypertens (Greenwich), 2022. 24(4): p. 438–448.

26. Hsing, S.C., et al., The Association of Losartan and Ramipril Therapy With Kidney and Cardiovascular Outcomes in Patients With Chronic Kidney Disease: A Chinese Nation-Wide Cohort Study in Taiwan. Medicine (Baltimore), 2015. 94(48): p. e1999.

27. Padwal, R., M. Lin, and D.T. Eurich, The Comparative Effectiveness of Angiotensin-Converting Enzyme Inhibitors and Angiotensin II Receptor Blockers in Patients With Diabetes. J Clin Hypertens (Greenwich), 2016. 18(3): p. 200–6.

28. Oger, E., L.-M. Scailteux, and S. Kerbrat, Comparative effectiveness of angiotensin-converting enzyme inhibitors and angiotensin receptor blockers on overall mortality and major cardiovascular diseases in primary prevention: Population-based nationwide cohort study based on French health insurance data (SNIIRAM)., in Abstracts of the 34th Inernational Conference on Pharmacoepidemiology & Therapeutic Risk Management. 2018: Prague Congress Centre, Prague, Czech Republic. p. 398.

29. Potier, L., et al., Angiotensin-converting enzyme inhibitors and angiotensin receptor blockers in high vascular risk. Heart, 2017. 103(17): p. 1339–1346.

30. Seminog, O.O., et al., Determinants of the decline in mortality from acute stroke in England: linked national database study of 795 869 adults. BMJ, 2019. 365: p. l1778.

