## Supplementary Table 1 for "Cardiorenal Effects of Angiotensin-converting enzyme inhibitors and Angiotensin receptor blockers in people underrepresented in trials: analysis of routinely collected data with validation against a target trial"

| **Supplementary Table 1.** Deviations from protocol | |
|---|---|
| **Deviation** | **Reason** |
| Using propensity-score—weighting as opposed to propensity-score—matching (with propensity-score—matching carried out as an additional analysis) | To obtain average treatment effect as opposed to average treatment effect on treated and increase sample size |
| Underrepresented group analysis on propensity-score—weighted sample as opposed to propensity-score—matched cohort | This was to increase sample size as comparison between both analyses gave almost identical results |
| Primary outcome: including both fatal and non-fatal events for stroke and myocardial infarction | Consistency with trial |
| Included additional outcome- main secondary outcome: composite of cardiovascular-related death, myocardial infarction or stroke | Consistency with trial |
| Angina inclusion criteria: Removed condition that needed to have previous coronary artery disease diagnosis | Misclassification |
| CABG Inclusion criteria: Removed condition that could be with angina | Only included events where CABG was within 4 years prior to avoid due to potential of capturing old events |
| Comparing reason for discontinuation to safety outcomes in trial as opposed to events occurring within 3 months | Consistency with safety outcomes reported in trial |
| Objective of extending follow-up for safety events | Have not yet addressed this objective due to difficulty replicating safety trial results |
| Renal function omitted from propensity-score model | Due to large amounts of missingness |
| Adherence assessed differently and instead reported proportions of patients receiving each drug at different timepoints | Consistency with trial |
| Additional subgroups studied | To further demonstrate trial replicability and quantify effect modification |
| On-treatment (per-protocol) analysis for secondary objectives 1 and 2 (extending findings to trial-underrepresented and excluded groups) | Not deemed necessary as on-treatment analysis was sufficiently comparable to ITT for primary outcome |
| Previously mentioned that patients had to meet inclusion and exclusion criteria prior to start of first exposed period instead trial criteria assessed at start of all exposed periods | Incorrect wording in protocol this reduces bias by assessing at start of follow up |
| Referred to analysis group as trial-analogous now analysis groups will be labelled as propensity-score—weighted trial-eligible for main analysis and propensity-score—matched trial-eligible for sensitivity analysis | To avoid confusion as only the ACEi trial-eligible cohort is trial-matched |
| Naming of nephropathy outcomes | Changed from nephropathy 1 and nephropathy 2 to loss of eGFR or ESKD and ESKD |
| Nephropathy 1 sensitivity analysis requiring 2 measurements at least 3 months apart for both eGFR<15 and 50% reduction in eGFR | Previously stated this is only required for 50% reduction in eGFR which was incorrect |
