## Supplementary Table 2 for "Cardiorenal Effects of Angiotensin-converting enzyme inhibitors and Angiotensin receptor blockers in people underrepresented in trials: analysis of routinely collected data with validation against a target trial"

| **Supplementary Table 2.** List of variables considered and included in propensity-score models 1 and 2 | | |
|---|---|---|
| **Variables included in propensity-score model** | **Model 1** | **Model 2** |
| Stroke/TIA | ✓ | ✓ |
| Peripheral artery disease | ✓ | ✓ |
| Coronary artery disease | ✓ | ✓ |
| Diabetes | ✓ | ✓ |
| High-risk diabetes | ✓ | ✓ |
| Age (years) | ✓ | ✓ |
| Sex | ✓ | ✓ |
| Ethnicity | ✓ | ✓ |
| BMI | ✓ | ✓ |
| SBP | ✓ | ✓ |
| DBP | ✓ | ✓ |
| Index of Multiple Deprivation (IMD) | | ✓ |
| Smoke status | ✓ | ✓ |
| Alcohol use | | ✓ |
| Statin use | | ✓ |
| Nitrate use | | ✓ |
| Diabetic treatment use | | ✓ |
| Diuretic use | | ✓ |
| CCB use | | ✓ |
| Betablocker use | | ✓ |
| Aspirin use | | ✓ |
| Antiplatelet use | | ✓ |
| Digoxin use | | |
| Anticoagulant use | | |
| Alphablocker use | | |
| No. of hospital admissions within 6 months prior | | ✓ |
| No. of GP appointments within 6 months prior | | ✓ |
| No. of medications within 6 months prior | | ✓ |
| Year of start of eligible period | | ✓ |
| Time since first eligible period (days) | | ✓ |
| No. of previous ACE inhibitor eligible periods | | ✓ |
| No. of previous ARB eligible periods | | ✓ |

Notes: TIA: transient ischaemic attack; BMI: body-mass index; SBP: systolic blood pressure; DBP: diastolic blood pressure.
Variables are measured at start of trial-eligible period or before.
Peripheral artery disease includes limb bypass surgery or angioplasty, limb/foot amputation or intermittent claudication.
Coronary artery disease includes previous MI, angina, coronary angioplasty or CABG.
SBP and DBP are measured within 6 months prior to start of trial-eligible period.
Medication use is within 3 months prior to start of trial-eligible period.
