## Supplementary Table 3 for "Cardiorenal Effects of Angiotensin-converting enzyme inhibitors and Angiotensin receptor blockers in people underrepresented in trials: analysis of routinely collected data with validation against a target trial"

**Supplementary Table 3.** Assessment of balance of variables included in propensity-score model for ARB vs ACEi analysis (M2) before and after weighting

| Characteristic | Before propensity-score—weighting | | | After propensity-score—weighting | | |
|---|---|---|---|---|---|---|
| | ACEi N=96,602 | ARB N=40,553 | Standardised difference | ACEi N=96,602 | ARB N=40,553 | Standardised difference |
| **Age - year** | 70.8 ± 9.0 | 71.2 ± 8.7 | 0.047 | 71.2 ± 8.9 | 71.1 ± 9.0 | 0.009 |
| **Blood pressure – mmHg** | 147.4 ± 20.7 / 80.1 ± 10.7 | 148.1 ± 20.7 / 79.7 ± 10.5 | 0.035 / 0.041 | 147.4 ± 20.3 / 79.5 ± 10.7 | 148.2 ± 20.8 / 80.1 ± 10.7 | 0.041 / 0.054 |
| **Body-mass index (kg/m$^2$)** | 28.3 ± 5.3 | 28.8 ± 5.4 | 0.097 | 28.3 ± 5.7 | 28.6 ± 5.4 | 0.044 |
| **Female sex – no. (%)** | 45508 (47.1) | 22690 (56.0) | 0.178 | 75304.7 (51.8) | 68530.9 (50.5) | 0.026 |
| **Ethnic group – no. (%)** | | | | | | |
| Black | 1280 (1.3) | 736 (1.8) | 0.039 | 2064.1 (1.4) | 2053.2 (1.5) | 0.008 |
| Other | 1134 (1.2) | 607 (1.5) | 0.028 | 1742.9 (1.2) | 1747.3 (1.3) | 0.008 |
| South Asian | 3026 (3.1) | 1799 (4.4) | 0.068 | 8632.0 (5.9) | 5073.8 (3.7) | 0.103 |
| White | 91162 (94.4) | 37411 (92.3) | 0.028 | 132818.6 (91.4) | 126718.0 (93.5) | 0.076 |
| **Clinical history – no. (%)** | | | | | | |
| CAD[1,2] | 68009 (70.4) | 28202 (69.5) | 0.019 | 100440.6 (69.2) | 94474.9 (69.7) | 0.011 |
| Cerebrovascular disease[1,3] | 8695 (9.0) | 3140 (7.7) | 0.046 | 12626.5 (8.7) | 11840.2 (8.7) | 0.001 |
| PAD[1,4] | 9999 (10.4) | 4078 (10.1) | 0.010 | 14359.7 (9.9) | 14463.3 (10.7) | 0.026 |
| Diabetes[5] | 43751 (45.3) | 20003 (49.3) | 0.081 | 69713.4 (48.0) | 64739.6 (47.8) | 0.005 |
| High-risk diabetes[1,6] | 30736 (31.8) | 14757 (36.4) | 0.097 | 50469.3 (34.7) | 46151.9 (34.0) | 0.015 |
| **Smoking status – no. (%)** | | | | | | |
| Non-smoker | 34503 (35.7) | 16470 (40.6) | 0.101 | 55849.7 (38.5) | 50717.2 (37.4) | 0.022 |

| **Supplementary Table 3.** Assessment of balance of variables included in propensity-score model for ARB vs ACEi analysis (M2) before and after weighting | | | | | | |
|---|---|---|---|---|---|---|
| **Characteristic** | **Before propensity-score—weighting** | | | **After propensity-score—weighting** | | |
| | ACEi N=96,602 | ARB N=40,553 | Standardised difference | ACEi N=96,602 | ARB N=40,553 | Standardised difference |
| Current smoker | 12921 (13.4) | 3552 (8.8) | 0.148 | 16659.7 (11.5) | 16627.5 (12.3) | 0.024 |
| Past smoker | 49178 (50.9) | 20531 (50.6) | 0.006 | 72748.3 (50.1) | 68247.6 (50.3) | 0.005 |
| **Alcohol status – no. (%)** | | | | | | |
| Drinker | 76756 (79.5) | 31840 (78.5) | 0.023 | 116186.3 (80.0) | 106324.0 (78.4) | 0.039 |
| **Medication[7] – no. (%)** | | | | | | |
| Antiplatelet agent | 12334 (12.8) | 2482 (6.1) | 0.229 | 14649.6 (10.1) | 13131.5 (9.7) | 0.014 |
| Aspirin | 44011 (45.6) | 9325 (23.0) | 0.489 | 53048.8 (36.5) | 50799.3 (37.5) | 0.020 |
| Beta-blocker | 34178 (35.4) | 6756 (16.7) | 0.437 | 40751.5 (28.1) | 38741.4 (28.6) | 0.012 |
| Calcium-channel blocker | 28820 (29.8) | 8515 (21.0) | 0.204 | 38254.1 (26.3) | 37864.7 (27.9) | 0.036 |
| Diuretics | 32002 (33.1) | 8838 (21.8) | 0.256 | 40789.2 (28.1) | 41887.5 (30.9) | 0.062 |
| Diabetic treatment | 20060 (20.8) | 4910 (12.1) | 0.235 | 25257.7 (17.4) | 25561.6 (18.9) | 0.038 |
| Nitrates | 14862 (15.4) | 3172 (7.8) | 0.238 | 17721.7 (12.2) | 16577.2 (12.2) | 0.001 |
| Statins | 52925 (54.8) | 11474 (28.3) | 0.558 | 64630.3 (44.5) | 61779.5 (45.6) | 0.021 |
| **Index of multiple deprivation (IMD) – no.(%)** | | | | | | |
| 1 (least deprived) | 19805 (20.5) | 8996 (22.2) | 0.041 | 32568.7 (22.4) | 28190.3 (20.8) | 0.040 |
| 2 | 21977 (22.8) | 9603 (23.7) | 0.022 | 34344.3 (23.6) | 30971.3 (22.8) | 0.019 |
| 3 | 20644 (21.4) | 8674 (21.4) | 0.001 | 30255.9 (20.8) | 28911.6 (21.3) | 0.012 |

**Supplementary Table 3.** Assessment of balance of variables included in propensity-score model for ARB vs ACEi analysis (M2) before and after weighting

| Characteristic | Before propensity-score—weighting | | | After propensity-score—weighting | | |
|---|---|---|---|---|---|---|
| | ACEi N=96,602 | ARB N=40,553 | Standardised difference | ACEi N=96,602 | ARB N=40,553 | Standardised difference |
| 4 | 18462 (19.1) | 7312 (18.0) | 0.028 | 26032.4 (17.9) | 25280.3 (18.6) | 0.019 |
| 5 (most deprived) | 15714 (16.3) | 5967 (14.7) | 0.043 | 22056.5 (15.2) | 22238.8 (16.4) | 0.033 |
| **Health utilisation**[8] | | | | | | |
| No. of hospital admissions | 0.42 ± 1.0 | 0.31 ± 0.9 | 0.118 | 0.38 ± 0.9 | 0.36 ± 0.9 | 0.017 |
| No. of GP apt. | 27.9 ± 27.3 | 15.2 ± 25.4 | 0.482 | 22.8 ± 26.9 | 24.3 ± 29.5 | 0.058 |
| No. of different drug types | 8.5 ± 4.3 | 9.7 ± 4.6 | 0.273 | 9.2 ± 4.5 | 9.0 ± 4.6 | 0.035 |
| **Time-related variables** | | | | | | |
| Time since first eligible period (days) | 131.3 ± 496.5 | 377.6 ± 759.1 | 0.384 | 544.7 ± 1444.4 | 223.6 ± 625.4 | 0.501 |
| No. of prior ARB periods | 0.04 ± 0.3 | 0.1 ± 0.5 | 0.163 | 0.1 ± 0.4 | 0.1 ± 0.4 | 0.117 |
| No. of prior ACEi periods | 0.1 ± 0.5 | 0.6 ± 0.7 | 0.739 | 0.3 ± 0.8 | 0.3 ± 0.6 | 0.053 |
| Calendar year | 2007.1 ± 4.0 | 2007.6 ± 4.1 | 0.131 | 2007.9 ± 4.6 | 2007.3 ± 4.1 | 0.152 |

Notes: Cohort includes 1 randomly selected eligible period per patient in each group
[1] Any diagnosis prior to start of eligible period
[2] Includes diagnosis of: MI at least 2 days prior, angina at least 30 days prior, angioplasty at least 30 days prior, CABG at least 4 years prior
[3] Includes diagnosis of: stroke/TIA
[4] Includes diagnosis of: limb bypass surgery, limb/foot amputation, intermittent claudication
[5] DM prior to start of eligible period
[6] Includes DM with: retinopathy, neuropathy, chronic kidney disease or proteinuria
[7] Within 3 months prior to eligible start date. Antiplatelet agent= clopidogrel/ ticlopidine
[8] Within 6 months prior to eligible start date.
no. (%)=number (percent); CAD=coronary artery disease; MI=myocardial infarction; PAD=peripheral artery disease.
