## Supplementary Table 4 for "Cardiorenal Effects of Angiotensin-converting enzyme inhibitors and Angiotensin receptor blockers in people underrepresented in trials: analysis of routinely collected data with validation against a target trial"

| **Supplementary Table 4.** Medication adherence to assigned exposure group | | | | |
|---|---|---|---|---|
| **Years of follow-up** | **ACEi patients** N=96,602 | | **ARB patients** N=40,553 | |
| | **Receiving** | | **Receiving** | |
| | **ACEi** | **ARB** | **ARB** | **ACEi** |
| 1 | 67347 (69.7) | 10805 (11.2) | 31661 (78.1) | 1039 (2.6) |
| 2 | 57188 (59.2) | 12177 (12.6) | 27381 (67.5) | 1483 (3.7) |
| 3 | 48991 (50.7) | 12148 (12.6) | 23487 (57.9) | 1718 (4.2) |
| 4 | 41583 (43.1) | 11523 (11.9) | 19950 (49.2) | 1828 (4.5) |
| 5.5 | 31769 (32.9) | 10060 (10.4) | 15210 (37.5) | 1764 (4.4) |
