## Supplementary Table 5 for "Cardiorenal Effects of Angiotensin-converting enzyme inhibitors and Angiotensin receptor blockers in people underrepresented in trials: analysis of routinely collected data with validation against a target trial"

**Supplementary Table 5.** Secondary and other outcomes after propensity-score—weighted analysis using CPRD data

| **Outcome** | **ACEi** (N=96,602) | **ARB** (N=40,553) | **ARB vs ACEi** |
|---|---|---|---|
| | *Number (percent)* | | *Hazard ratio (95% CI)* |
| Newly diagnosed congestive heart failure | 10232 (10.6) | 4017 (9.9) | 0.99 (0.94, 1.04) |
| Revascularisation procedures | 14132 (14.6) | 5250 (13.0) | 1.00 (0.96, 1.04) |
| Loss of GFR or ESKD | 4217 (5.2) | 2205 (6.1) | 1.11 (1.04, 1.19) |
| ESKD | 1460 (1.8) | 822 (2.3) | 1.06 (0.95, 1.19) |
| Microvascular complications of diabetes mellitus | 2261 (17.4) | 757 (14.4) | 0.95 (0.85, 1.05) |

Notes: ESKD: end-stage kidney disease.
CPRD weighted analysis includes 1 randomly selected eligible period per patient. Propensity-score—weighted with robust standard errors. ARB vs ACEi also adjusted for time since first eligible period, calendar year and number of prior ARB periods.
Loss of GFR or ESKD defined as: 50% reduction in estimated glomerular filtration ratio (eGFR), start of kidney replacement therapy (KRT) or eGFR<15.
ESKD defined as: start of KRT or eGFR<15.
Kidney outcomes only include those subjects who have an eGFR measurement before start of the eligible period but within 6 months and also adjusted for baseline serum creatinine.
Microvascular complications of diabetes mellitus outcome only include patients who are diabetic but non-high risk.
