## Supplementary Table 6 for "Cardiorenal Effects of Angiotensin-converting enzyme inhibitors and Angiotensin receptor blockers in people underrepresented in trials: analysis of routinely collected data with validation against a target trial"

**Supplementary Table 6.** Reason for treatment cessation using trial criteria and propensity-score—weighting for ARB vs ACEi compared to ONTARGET

| Reason for cessation | CPRD | | | ONTARGET |
|---|---|---|---|---|
| | **ARB** (N=40,553) | **ACEi** (N=96,602) | **ARB vs ACEi** | **telmisartan vs ramipril** |
| | *Number (percent)* | | *Relative risk (95% CI)* | *Relative risk (P value)* |
| Cough | 949 (2.3) | 1557 (1.6) | 1.29 (1.16, 1.43) | 0.26 (<0.001) |
| Angioedema | 37 (0.09) | 83 (0.09) | 1.14 (0.72, 1.80) | 0.4 (0.01) |
| Hyperkalaemia[1] | 2784 (7.8) | 5836 (7.4) | 1.12 (1.06, 1.18) | |
| ≥30% increase in serum creatinine | 7222 (19.8) | 12441 (15.2) | 1.38 (1.34, 1.43) | 1.14 (0.46)[2] |

Notes: In CPRD, treatment cessation is defined as the end date of the trial-eligible exposed period included in analysis (i.e., the date prior to a prescription gap of >90 days) and the latest event occurring prior to end of trial-eligible period is counted as the reason for treatment cessation. Multiple reasons that occur on the same day are both counted.
Analysis is adjusted for time since first eligible period, calendar year and number of prior ARB eligible periods.
[1]Defined as potassium >5.5 mmol/l. Analysis out of number of people with non-missing potassium.
Definition of renal impairment as reason for discontinuation in ONTARGET is not stated so results are not directly comparable to CPRD
Kidney outcomes are adjusted for baseline serum creatinine and are out of the number of people with non-missing eGFR in CPRD.
ONTARGET did not present 95% CI.
