## Supplementary Table 7 for "Cardiorenal Effects of Angiotensin-converting enzyme inhibitors and Angiotensin receptor blockers in people underrepresented in trials: analysis of routinely collected data with validation against a target trial"

**Supplementary Table 7.** Number of events in the primary outcome, its components, and death from any cause for ARB vs ACEi using a propensity-score—matched analysis of patients in CPRD (sensitivity analysis)

| **Outcome** | **CPRD: Propensity-score—matched** | | | **ONTARGET** |
|---|---|---|---|---|
| | **ACEi** (N=15,462) | **ARB** (N=15,462) | **ARB vs ACEi** | **Telmisartan vs Ramipril** |
| | *Number (percent)* | | *Hazard ratio (95% CI)* | |
| Primary composite: Death from cardiovascular causes, myocardial infarction, stroke, or hospitalisation for heart failure | 2539 (16.4) | 2453 (15.9) | 0.97 (0.92, 1.02) | 1.01 (0.94, 1.09) |
| Main secondary outcome: Death from cardiovascular causes, myocardial infarction or stroke | 2234 (14.5) | 2173 (14.1) | 0.98 (0.92, 1.04) | 0.99 (0.91, 1.07) |
| Myocardial infarction | 1806 (11.7) | 1721 (11.1) | 0.96 (0.90, 1.03) | 1.07 (0.94, 1.22) |
| Stroke | 535 (3.5) | 591 (3.8) | 1.10 (0.98, 1.24) | 0.91 (0.79, 1.05) |
| Hospitalisation for heart failure | 542 (3.5) | 513 (3.3) | 0.94 (0.83, 1.06) | 1.12 (0.97, 1.29) |
| Death from cardiovascular causes | 655 (4.2) | 649 (4.2) | 0.98 (0.88, 1.09) | 1.00 (0.89, 1.12) |
| Death from non-cardiovascular causes | 852 (5.5) | 856 (5.5) | 0.99 (0.90, 1.09) | 0.96 (0.83, 1.10) |
| Death from any cause | 1507 (9.8) | 1505 (9.7) | 0.99 (0.92, 1.06) | 0.98 (0.90, 1.07) |

Notes: Propensity-score—matched cohort developed using trial-matched ACEi patients 1:1 matched to closest trial-eligible ARB period.
Myocardial infarction and stroke include both fatal and non-fatal events.
ONTARGET results are from published findings.
