## Supplementary Table 8 for "Cardiorenal Effects of Angiotensin-converting enzyme inhibitors and Angiotensin receptor blockers in people underrepresented in trials: analysis of routinely collected data with validation against a target trial"

| **Supplementary Table 8:** Safety outcomes assessed among non-switchers using trial criteria and propensity-score—weighting for ARB vs ACEi (sensitivity analysis) | | | | |
|---|---|---|---|---|
| **Safety outcome** | **CPRD** | | | **ONTARGET** |
| | **Reason for treatment cessation** | | | |
| | **ARB** (N=11,856) | **ACEi** (N=90,597) | **ARB vs ACEi** | **telmisartan vs ramipril** |
| | *Number (percent)* | | *Relative risk (95% CI)* | *Relative risk (P value)* |
| Cough | 178 (1.5) | 1455 (1.6) | 0.84 (0.70, 1.01) | 0.26 (<0.001) |
| Angioedema | 10 (0.08) | 77 (0.08) | 0.72 (0.35, 1.48) | 0.4 (0.01) |
| Hyperkalaemia[1] | 685 (7.7) | 5473 (7.4) | 1.06 (0.97, 1.16) | |
| ≥30% increase in serum creatinine | 1730 (18.8) | 11781 (15.5) | 1.25 (1.18, 1.31) | 1.14 (0.01)[2] |

Notes: In CPRD, treatment cessation is defined as the end date of the trial-eligible exposed period included in analysis (i.e., the date prior to a prescription gap of >90 days) and the latest event occurring prior to end of trial-eligible period is counted as the reason for treatment cessation. Multiple reasons that occur on the same day are both counted.

Reason for treatment cessation represents the main analysis which is compared to ONTARGET and events occurring within 3 months represents the additional analysis exploring safety events which occur within 3 months of the start of eligible period.

Non-switchers include first trial-eligible period per patient and excludes patients with previous exposure to opposing drug at any time prior to start of included trial-eligible period.

Both analyses are adjusted for number of previous GP appointments within 6 months prior.

[1]Defined as potassium >5.5 mmol/l. Analysis out of number of people with non-missing potassium.

Definition of kidney impairment in ONTARGET is not stated so results are not directly comparable to CPRD

Kidney outcomes are adjusted for baseline serum creatinine and are out of the number of people with non-missing eGFR in CPRD.

ONTARGET did not present 95% CI.
