## Supplementary Figure 1 for "Cardiorenal Effects of Angiotensin-converting enzyme inhibitors and Angiotensin receptor blockers in people underrepresented in trials: analysis of routinely collected data with validation against a target trial"

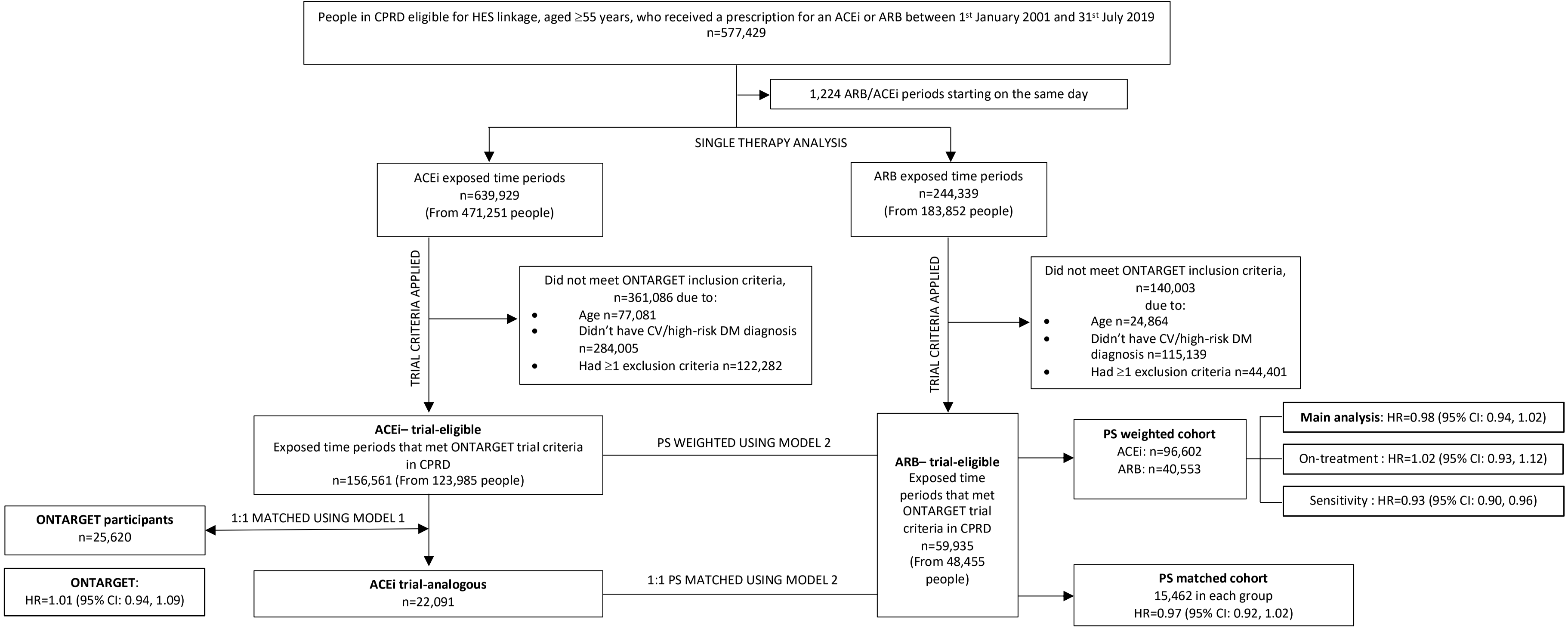


**Supplementary Figure 1. Single therapy study profile**

DM: diabetes mellitus; PS: propensity score. Model 1 is the propensity score model for the probability of being included in the trial. Model 2 is the propensity score model for probability of receiving an ACEi built using the cohort of ACEi trial-analogous patients appended to the ARB trial-eligible patients. The PS weighted analysis uses inverse PS weights generated from running Model 2 on 1 randomly selected trial-eligible ARB period per patient and 1 randomly selected trial-eligible ACEi period per patient. For the main analysis follow-up was from the start date of the trial-eligible period and patients were censored at the earliest date of: outcome, death, transferred out of practice, last practice data collection, or 5.5 years from the start of trial-eligible period, to reflect the maximum follow-up in the trial. In the on-treatment analysis, patients were additionally censored at the end of an eligible period, if they switched treatment or started dual therapy. This was denoted as date of last drug and patients were censored at this date +60 days. In the sensitivity analysis follow-up started from 28 days after the start of the eligible period (to reflect the typical length of prescription), excluding patients if there were no prescriptions after 28 days. 51,775 ACEi patients and 18,410 ARB patients were excluded for heart failure.
