## Supplementary Figure 2 for "Cardiorenal Effects of Angiotensin-converting enzyme inhibitors and Angiotensin receptor blockers in people underrepresented in trials: analysis of routinely collected data with validation against a target trial"

| Outcome | ARB (N=40,553) n (%) | ACEi (N=96,602) n (%) | ARB vs ACEi HR (95% CI) | P Value for Interaction |
|---|---|---|---|---|
| Primary composite | | | | 0.21 |
| Male | 3166 (17.7) | 9931 (19.4) | 0.94 (0.88, 1.00) | |
| Female | 3121 (13.8) | 7004 (15.4) | 1.00 (0.94, 1.06) | |
| Main secondary | | | | 0.37 |
| Male | 2746 (15.4) | 8747 (17.1) | 0.95 (0.89, 1.01) | |
| Female | 2617 (11.5) | 5900 (13.0) | 1.00 (0.93, 1.07) | |
| Myocardial infarction | | | | 0.25 |
| Male | 2228 (12.4) | 7344 (14.4) | 0.92 (0.86, 0.99) | |
| Female | 1862 ( 8.2) | 4273 ( 9.4) | 0.99 (0.91, 1.08) | |
| Stroke | | | | 0.14 |
| Male | 665 ( 3.7) | 1811 ( 3.5) | 1.10 (0.98, 1.23) | |
| Female | 908 ( 4.0) | 1957 ( 4.3) | 0.98 (0.89, 1.08) | |
| Hospitalisation for HF | | | | 0.81 |
| Male | 761 ( 4.3) | 2158 ( 4.2) | 0.96 (0.86, 1.07) | |
| Female | 809 ( 3.6) | 1870 ( 4.1) | 0.94 (0.84, 1.05) | |
| Death from CV causes | | | | 0.03 |
| Male | 897 ( 5.0) | 2791 ( 5.5) | 1.02 (0.93, 1.12) | |
| Female | 928 ( 4.1) | 2403 ( 5.3) | 0.88 (0.80, 0.97) | |
| Death from non-CV causes | | | | 0.32 |
| Male | 1223 ( 6.8) | 3665 ( 7.2) | 0.98 (0.91, 1.07) | |
| Female | 1426 ( 6.3) | 3319 ( 7.3) | 0.93 (0.86, 1.00) | |
| Death from any cause | | | | 0.04 |
| Male | 2120 (11.9) | 6456 (12.6) | 1.00 (0.94, 1.06) | |
| Female | 2354 (10.4) | 5722 (12.6) | 0.91 (0.85, 0.97) | |
| Newly diagnosed HF | | | | 0.62 |
| Male | 1944 (10.9) | 5649 (11.1) | 0.96 (0.90, 1.02) | |
| Female | 2073 ( 9.1) | 4583 (10.1) | 0.98 (0.92, 1.05) | |
| Revascularisation proc | | | | 0.03 |
| Male | 3397 (19.0) | 10185 (19.9) | 0.96 (0.91, 1.01) | |
| Female | 1853 ( 8.2) | 3947 ( 8.7) | 1.06 (0.98, 1.15) | |
| Loss of GFR or ESKD | | | | 0.32 |
| Male | 965 ( 6.0) | 2026 ( 4.8) | 1.13 (1.03, 1.25) | |
| Female | 1240 ( 6.1) | 2191 ( 5.5) | 1.06 (0.97, 1.16) | |
| ESKD | | | | 0.79 |
| Male | 380 ( 2.4) | 718 ( 1.7) | 1.06 (0.90, 1.25) | |
| Female | 442 ( 2.2) | 742 ( 1.9) | 1.03 (0.88, 1.20) | |
| Microvascular comps of DM | | | | 0.80 |
| Male | 428 (17.1) | 1455 (19.6) | 0.92 (0.80, 1.06) | |
| Female | 329 (12.0) | 806 (14.4) | 0.95 (0.80, 1.12) | |

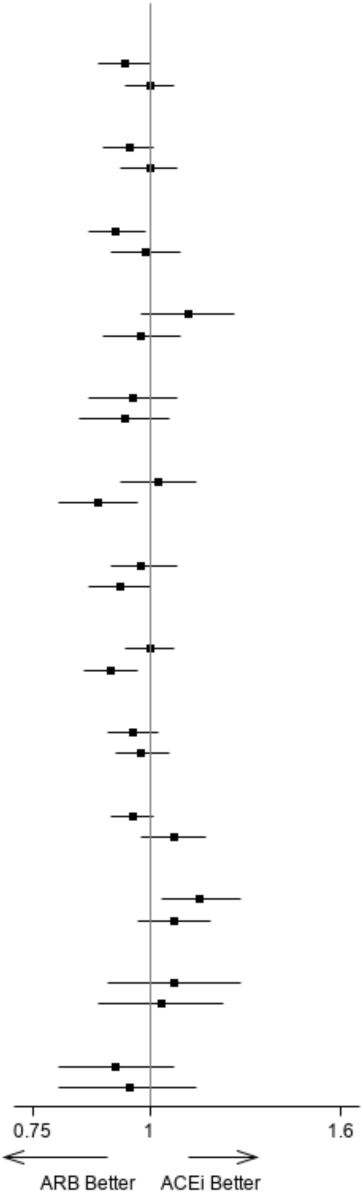


**Supplementary Figure 2.** Treatment heterogeneity by sex for all outcomes for comparison of ARB vs ACEi. n (%)= number of events (percent). P-value is the test of interaction between the treatment for each outcome. ESKD: end-stage kidney disease. Analysis is propensity-score—weighted with robust standard errors. Analysis adjusted for number of previous GP appointments and medications within 6 months prior, time since first eligible period and number of prior ARB periods. Loss of GFR or ESKD defined as: 50% reduction in estimated glomerular filtration rate (GFR), start of kidney replacement therapy (KRT) or eGFR<15. ESKD defined as: start of KRT or eGFR<15. Kidney outcomes only include those subjects who have an eGFR measurement before start of the eligible period but within 6 months and also adjusted for baseline serum creatinine. Microvascular complications of diabetes mellitus outcome only include patients who are diabetic but non-high risk.
