## Supplementary Figure 3 for "Cardiorenal Effects of Angiotensin-converting enzyme inhibitors and Angiotensin receptor blockers in people underrepresented in trials: analysis of routinely collected data with validation against a target trial"

| Outcome | ARB (N=36,919) n (%) | ACEi (N=83,515) n (%) | ARB vs ACEi HR (95% CI) | P Value for Interaction |
|---|---|---|---|---|
| Primary composite | | | | 0.62 |
| <65 yr | 1127 (11.1) | 3625 (13.4) | 0.99 (0.91, 1.08) | |
| >=65 to <75 yr | 2096 (13.6) | 5489 (15.7) | 0.97 (0.90, 1.05) | |
| >=75 yr | 3064 (20.3) | 7821 (22.6) | 0.94 (0.89, 1.00) | |
| Main secondary | | | | 0.64 |
| <65 yr | 1023 (10.1) | 3350 (12.3) | 1.00 (0.92, 1.10) | |
| >=65 to <75 yr | 1809 (11.8) | 4831 (13.8) | 0.97 (0.89, 1.05) | |
| >=75 yr | 2531 (16.8) | 6466 (18.7) | 0.95 (0.89, 1.02) | |
| Myocardial infarction | | | | 0.73 |
| <65 yr | 870 ( 8.6) | 2973 (11.0) | 0.99 (0.89, 1.09) | |
| >=65 to <75 yr | 1413 ( 9.2) | 3922 (11.2) | 0.95 (0.86, 1.05) | |
| ≥75 yr | 1807 (12.0) | 4722 (13.7) | 0.94 (0.86, 1.02) | |
| Stroke | | | | 0.45 |
| <65 yr | 188 ( 1.9) | 478 ( 1.8) | 1.16 (0.94, 1.43) | |
| >=65 to <75 yr | 494 ( 3.2) | 1142 ( 3.3) | 1.01 (0.88, 1.16) | |
| >=75 yr | 891 ( 5.9) | 2148 ( 6.2) | 1.00 (0.91, 1.10) | |
| Hospitalisation for HF | | | | 0.79 |
| <65 yr | 187 ( 1.8) | 489 ( 1.8) | 0.99 (0.78, 1.25) | |
| >=65 to <75 yr | 486 ( 3.2) | 1201 ( 3.4) | 0.97 (0.85, 1.11) | |
| >=75 yr | 897 ( 5.9) | 2338 ( 6.8) | 0.93 (0.84, 1.02) | |
| Death from CV causes | | | | 0.89 |
| <65 yr | 153 ( 1.5) | 455 ( 1.7) | 0.99 (0.79, 1.25) | |
| >=65 to <75 yr | 454 ( 3.0) | 1275 ( 3.7) | 0.93 (0.81, 1.06) | |
| >=75 yr | 1218 ( 8.1) | 3464 (10.0) | 0.94 (0.87, 1.02) | |
| Death from non-CV causes | | | | 0.23 |
| <65 yr | 261 ( 2.6) | 669 ( 2.5) | 1.07 (0.90, 1.28) | |
| >=65 to <75 yr | 803 ( 5.2) | 1969 ( 5.6) | 0.97 (0.88, 1.08) | |
| >=75 yr | 1585 (10.5) | 4346 (12.6) | 0.91 (0.85, 0.99) | |
| Death from any cause | | | | 0.32 |
| <65 yr | 414 ( 4.1) | 1124 ( 4.1) | 1.04 (0.90, 1.20) | |
| >=65 to <75 yr | 1257 ( 8.2) | 3244 ( 9.3) | 0.96 (0.88, 1.04) | |
| >=75 yr | 2803 (18.6) | 7810 (22.6) | 0.93 (0.88, 0.98) | |
| Newly diagnosed HF | | | | 0.50 |
| <65 yr | 505 ( 5.0) | 1494 ( 5.5) | 0.91 (0.79, 1.04) | |
| >=65 to <75 yr | 1276 ( 8.3) | 3171 ( 9.1) | 0.96 (0.88, 1.04) | |
| >=75 yr | 2236 (14.8) | 5567 (16.1) | 0.99 (0.93, 1.05) | |
| Revascularisation proc | | | | 0.02 |
| <65 yr | 1373 (13.6) | 4669 (17.2) | 0.91 (0.84, 0.98) | |
| >=65 to <75 yr | 2219 (14.4) | 5637 (16.1) | 1.04 (0.97, 1.12) | |
| >=75 yr | 1658 (11.0) | 3826 (11.1) | 1.03 (0.95, 1.12) | |
| Loss of GFR or ESKD | | | | 0.00 |
| <65 yr | 434 ( 4.8) | 675 ( 3.1) | 1.42 (1.22, 1.66) | |
| >=65 to <75 yr | 745 ( 5.4) | 1426 ( 4.8) | 1.09 (0.97, 1.22) | |
| >=75 yr | 1026 ( 7.5) | 2116 ( 7.0) | 1.00 (0.91, 1.10) | |
| ESKD | | | | 0.06 |
| <65 yr | 197 ( 2.2) | 266 ( 1.2) | 1.28 (1.00, 1.63) | |
| >=65 to <75 yr | 291 ( 2.1) | 495 ( 1.7) | 1.12 (0.92, 1.37) | |
| >=75 yr | 334 ( 2.4) | 699 ( 2.3) | 0.91 (0.77, 1.07) | |
| Microvascular comps of DM | | | | 0.24 |
| <65 yr | 282 (16.6) | 921 (19.9) | 0.96 (0.81, 1.13) | |
| >=65 to <75 yr | 352 (15.6) | 954 (17.8) | 0.99 (0.84, 1.15) | |
| >=75 yr | 123 ( 9.5) | 386 (12.7) | 0.76 (0.59, 0.99) | |

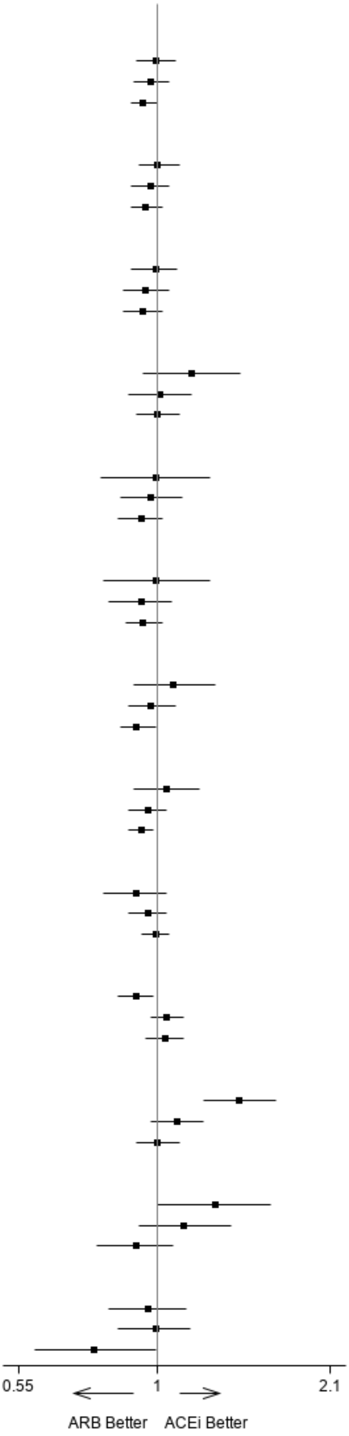


**Supplementary Figure 3.** Treatment heterogeneity by age group for all outcomes for comparison of ARB vs ACEi.

n (%)= number of events (percent). P-value is the test of interaction between the treatment for each outcome. ESKD: end-stage kidney disease. Analysis is propensity-score—weighted with robust standard errors. Analysis adjusted for number of previous medications within 6 months prior, time since first eligible period, number of prior ARB and ACEi periods, age and DBP. Loss of GFR or ESKD defined as: 50% reduction in estimated glomerular filtration rate (eGFR), start of kidney replacement therapy (KRT) or eGFR<15. ESKD defined as: start of KRT or eGFR<15. Kidney outcomes only include those subjects who have an eGFR measurement before start of the eligible period but within 6 months and also adjusted for baseline serum creatinine. Microvascular complications of diabetes mellitus outcome only include patients who are diabetic but non-high risk.
