## Supplementary Figure 4 for "Cardiorenal Effects of Angiotensin-converting enzyme inhibitors and Angiotensin receptor blockers in people underrepresented in trials: analysis of routinely collected data with validation against a target trial"

| Outcome | ARB (N=36,919) n (%) | ACEi (N=83,515) n (%) | ARB vs ACEi HR (95% CI) | P Value for Interaction |
|---|---|---|---|---|
| Primary composite | | | | 0.17 |
| No CKD | 3126 (13.3) | 8417 (15.2) | 0.94 (0.89, 0.99) | |
| CKD | 2484 (18.4) | 5553 (19.6) | 1.00 (0.94, 1.06) | |
| Main secondary | | | | 0.06 |
| No CKD | 2716 (11.6) | 7405 (13.4) | 0.94 (0.88, 1.00) | |
| CKD | 2058 (15.3) | 4556 (16.1) | 1.03 (0.96, 1.10) | |
| Myocardial infarction | | | | 0.02 |
| No CKD | 2139 ( 9.1) | 5973 (10.8) | 0.92 (0.86, 0.99) | |
| CKD | 1482 (11.0) | 3310 (11.7) | 1.05 (0.97, 1.14) | |
| Stroke | | | | 0.67 |
| No CKD | 709 ( 3.0) | 1782 ( 3.2) | 0.97 (0.86, 1.09) | |
| CKD | 705 ( 5.2) | 1516 ( 5.4) | 1.00 (0.90, 1.12) | |
| Hospitalisation for HF | | | | 0.68 |
| No CKD | 676 ( 2.9) | 1758 ( 3.2) | 0.91 (0.80, 1.02) | |
| CKD | 740 ( 5.5) | 1694 ( 6.0) | 0.94 (0.84, 1.05) | |
| Death from CV causes | | | | 0.21 |
| No CKD | 697 ( 3.0) | 2109 ( 3.8) | 0.87 (0.78, 0.97) | |
| CKD | 966 ( 7.2) | 2464 ( 8.7) | 0.96 (0.87, 1.05) | |
| Death from non-CV causes | | | | 0.81 |
| No CKD | 1152 ( 4.9) | 3159 ( 5.7) | 0.95 (0.87, 1.03) | |
| CKD | 1290 ( 9.6) | 3065 (10.8) | 0.96 (0.88, 1.04) | |
| Death from any cause | | | | 0.33 |
| No CKD | 1849 ( 7.9) | 5268 ( 9.5) | 0.92 (0.85, 0.98) | |
| CKD | 2256 (16.7) | 5529 (19.5) | 0.96 (0.90, 1.02) | |
| Newly diagnosed HF | | | | 0.03 |
| No CKD | 1774 ( 7.6) | 4798 ( 8.7) | 0.91 (0.84, 0.98) | |
| CKD | 1859 (13.8) | 4037 (14.3) | 1.02 (0.95, 1.10) | |
| Revascularisation proc | | | | 0.01 |
| No CKD | 3096 (13.2) | 8370 (15.2) | 0.95 (0.89, 1.00) | |
| CKD | 1616 (12.0) | 3145 (11.1) | 1.08 (0.99, 1.17) | |
| Loss of GFR or ESKD | | | | 0.85 |
| No CKD | 945 ( 4.0) | 2047 ( 3.7) | 1.11 (1.00, 1.23) | |
| CKD | 1404 (10.4) | 2420 ( 8.5) | 1.10 (0.99, 1.21) | |
| ESKD | | | | 0.39 |
| No CKD | 176 ( 0.8) | 418 ( 0.8) | 1.00 (0.80, 1.25) | |
| CKD | 789 ( 5.8) | 1253 ( 4.4) | 1.13 (0.96, 1.32) | |

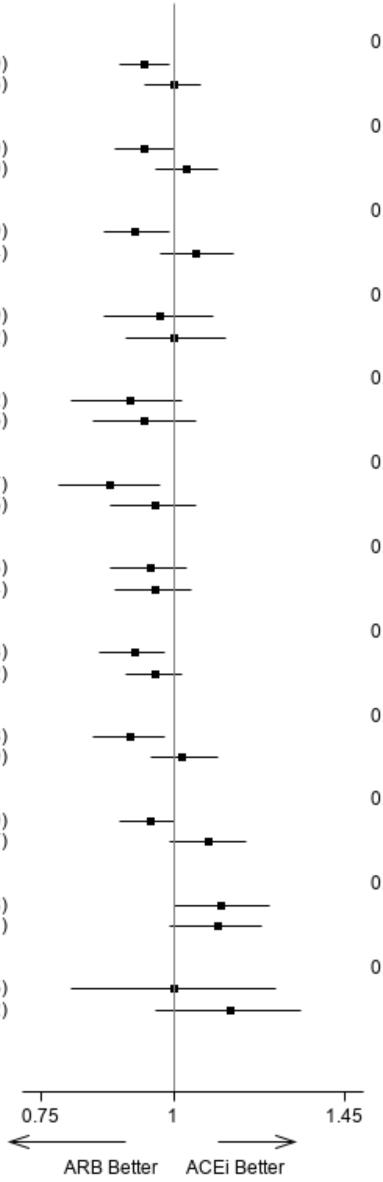


**Supplementary Figure 4.** Treatment heterogeneity by CKD status for all outcomes for comparison of ARB vs ACEi.

CKD: estimated GFR <60 mL/min/1.73m$^2$; ESKD: end-stage kidney disease. n (%)= number of events (percent). P-value is the test of interaction between the treatment for each outcome. Analysis is propensity-score—weighted with robust standard errors. Analysis adjusted for time since first eligible period. Loss of GFR or ESKD defined as: 50% reduction in estimated glomerular filtration rate (eGFR), start of kidney replacement therapy (KRT) or eGFR<15. ESKD defined as: start of KRT or eGFR<15. Kidney outcomes only include those subjects who have an eGFR measurement before start of the eligible period but within 6 months and also adjusted for baseline serum creatinine.
